# Rapid magnetic bead nucleic acid extraction enhances influenza RT-qPCR sensitivity and subtyping success

**DOI:** 10.64898/2026.08.18.26360610

**Authors:** Matthew L. Cavuto, Suleyman Sarp Pinar, Javier Sánchez-Martínez, Carla Rodríguez-Crespo, Ivana Pennisi, Katarzyna Szostak-Lipowicz, Nicolas Moser, Kenny Malpartida-Cardenas, Alison H Holmes, José María Eiros, Jesus Rodriguez-Manzano, Iván Sanz-Muñoz

**Author notes:** These authors contributed equally (Matthew L. Cavuto, Suleyman Sarp Pinar). Both authors should be considered senior authors (Jesus Rodriguez-Manzano, Iván Sanz-Muñoz).

## Abstract

Nucleic acid extraction remains the principal infrastructure barrier to molecular influenza testing outside centralised laboratories, since bead-based purification is normally tied to mains-powered extractors and trained operators. We evaluated SmartLid, a centrifugation-free format in which a removable magnetic key shuttles paramagnetic beads through pre-aliquoted lysis/binding, wash, and elution buffers without pipetting or powered instrumentation, against an automated magnetic-bead extractor (Nextractor NX-48S) on 311 nasopharyngeal specimens from the 2024-2025 influenza season at a National Influenza Centre. Paired eluates were amplified under identical monoplex RT-qPCR conditions for influenza A(H1N1)pdm09, A(H3), and B/Victoria. Both methods gave 100% specificity (47/47 negatives; no false positives). Subtyping succeeded in 263/264 reference-positive specimens after SmartLid extraction versus 241/264 after automated extraction (99.62% versus 91.29%; difference 8.33 percentage points; discordant pairs 23 versus 1; McNemar P < 0.001). Across 240 complete pairs, cycle threshold (Ct) values were lower after SmartLid extraction (median paired difference −2.78 cycles; estimated location shift −2.60 cycles, 95% CI −2.82 to −2.37; P < 0.001) with rank-ordering of specimens conserved between methods (Spearman rho = 0.84). The advantage was preserved across all three subtypes and in both fresh and frozen specimens (adjusted P < 0.001). Specimens recovered only after SmartLid extraction had higher Ct values than dual-detected specimens (median 34.37 versus 28.54; P < 0.001), locating the gain near the assay detection limit. An instrument-free manual format can therefore exceed the extraction efficiency of an automated comparator workflow, extending quality-assured influenza subtyping beyond centralised laboratories.

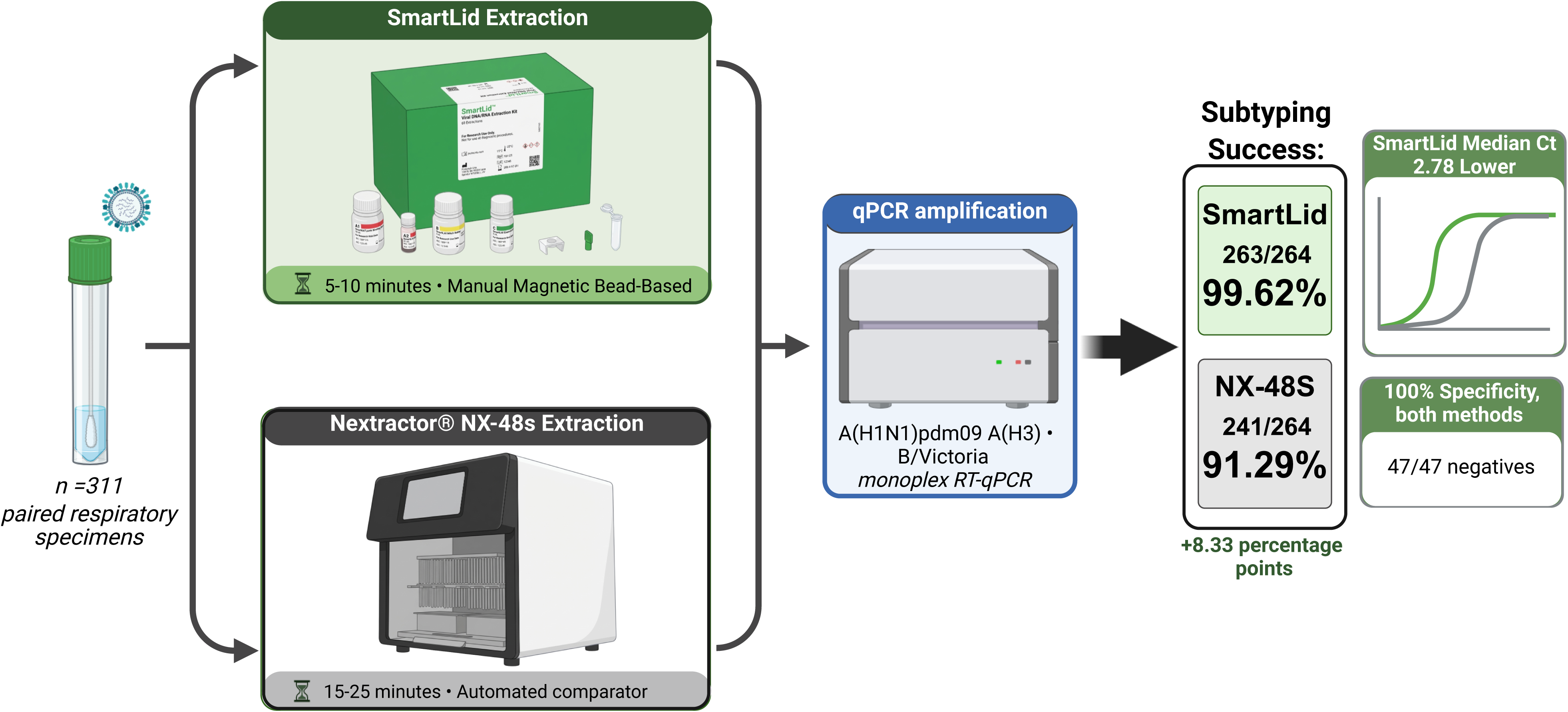

## INTRODUCTION

Influenza imposes a substantial clinical and economic burden worldwide (1,2). In Spain, for example, multi-season analyses from national surveillance and hospital datasets indicate roughly 29,000 influenza-related hospitalisations, 2,200 intensive care unit (ICU) admissions, and 1,600 in-hospital deaths each flu season; across the last five seasons, 127,160 hospitalizations were recorded, with a 7.4% ICU admission rate and 5.8% case fatality (3,4). More broadly, influenza causes an estimated 15,000–70,000 deaths annually in Europe and 290,000– 650,000 respiratory deaths worldwide each year (2). Together, these figures underscore the need for timely, sensitive diagnostics and robust surveillance to reduce morbidity and mortality.

A major limitation in diagnosing influenza is the continued reliance on syndromic identification of influenza-like illness (ILI). ILI case definitions are useful for surveillance but are not meant to give a final clinical diagnosis, as influenza is difficult to distinguish clinically from other respiratory diseases (5,6). This limits their utility in patient management. Recently released results showed a high sensitivity of ILI for diagnosing influenza but a low specificity, suggesting that many patients who meet ILI criteria may not have laboratory-confirmed influenza (5). Molecular diagnostics for influenza rely on centralised microbiological laboratories and molecular assays, which can be costly, time-consuming, and dependent on qualified individuals and laboratory equipment (7). In primary care, these delays could lead to significantly delayed result turnaround times, which are critical for guiding treatment decisions, such as antiviral use, antibiotic stewardship, and patient management. Therefore, there is a need for economical, reliable, fast, and easy-to-use point-of-care tools for both clinical decision-making and influenza surveillance.

Accurate influenza detection and subtyping rely on efficient nucleic acid extraction for downstream molecular analysis (8). However, this step generally depends on laboratory equipment, including automated extractors, that requires mains power, fixed infrastructure, and trained staff, constraining molecular testing outside centralized laboratories (9). Such constraints have driven interest in simplified, more accessible sample-preparation methods suitable for deployment at or near the point of care (POC) (10,11).

Accordingly, this study assessed whether a centrifugation-free nucleic acid extraction method, SmartLid (12–14), could support accurate downstream influenza detection and subtyping, using an automated extraction workflow as the gold-standard comparator. The SmartLid technology employs a custom magnetic lid to shuttle magnetic beads through lysis/binding, wash, and elution steps, enabling efficient nucleic acid extraction without requiring powered instrumentation or specialised training (12). A removable magnetic key integrated into SmartLid captures and releases paramagnetic beads, shuttling them sequentially through pre-aliquoted lysis/binding, wash, and elution buffers without pipetting or powered instrumentation. The system was initially engineered as a single-use, fully self-contained point-of-care (POC) extraction kit to enable rapid, electricity-free nucleic acid purification in low-resource settings; however, subsequent optimisation led to the development of a higher-throughput bulk- format capable of processing up to 12 samples simultaneously while preserving the same magnetic shuttling principle (14). Whereas SmartLid was previously validated for SARS-CoV-2 detection against a manual spin-column reference (12), here we benchmark it against a fully automated magnetic-bead extractor and extend the read-out from detection to influenza subtyping, providing a stringent, clinically relevant test of extraction efficiency under routine public-health laboratory conditions.

This study was conducted at the National Influenza Centre of Valladolid (NICV), Spain, where 311 clinical respiratory specimens were processed in parallel by SmartLid and an established automated magnetic-bead workflow (4,15,16), followed by downstream amplification and subtyping by RT-qPCR. Across these 311 specimens, SmartLid matched the automated comparator’s perfect specificity while achieving higher subtyping success, yielded lower Cts with preserved rank ordering across subtypes and storage conditions, and recovered additional low-load samples missed by the comparator—together pointing to greater analytical sensitivity in the downstream workflow.

## METHODS

### Specimens and classification

An observational analytical study was conducted at the National Influenza Centre of Valladolid using deidentified nasopharyngeal swab specimens collected during the 2024–2025 influenza season as part of routine acute respiratory infection and severe acute respiratory infection surveillance in Castilla y León, Spain. Specimens were obtained through sentinel primary-care facilities, non-sentinel primary-care centres, and regional hospitals, following the surveillance workflow previously described by Sanz-Muñoz et al. (15).

Nasopharyngeal specimens were collected using sterile nylon microfibre swabs supplied in 2 mL transport medium (TM014; Vircell S.L., Spain). Reference influenza status was established before the comparative extraction study using the site’s routine diagnostic workflow, which included the FilmArray® Respiratory Panel (BioFire Diagnostics) and influenza RT-qPCR testing (15,17).

A total of 311 clinical specimens were included. Of these, 264/311 (84.9%) were classified as influenza-positive and represented influenza A(H1N1)pdm09, influenza A(H3), or influenza B/Victoria; the remaining 47/311 (15.1%) were influenza-negative and served as negative controls. Among the 264 influenza-positive specimens, 162 were analysed fresh and 102 had been stored at ≤−70°C before extraction. Each specimen was processed in parallel using the manual SmartLid Viral DNA/RNA Extraction method and the automated Nextractor® NX-48S workflow.

### Nucleic acid extraction

Clinical specimens were extracted in parallel using two magnetic bead-based workflows. For automated extraction, specimens were vortexed and 200 µL of each sample was transferred to the extraction plate. Nucleic acids were purified using the RNA/DNA Pathogen Extraction Kit (reference MAD-003955M-EX-A24; Vitro Master Diagnóstica, Spain) on the Nextractor® NX-48S platform (Genolution, Republic of Korea), using the manufacturer’s VN protocol. The final eluate volume was approximately 20 µL.

In parallel, all samples were extracted with the SmartLid Viral DNA/RNA Extraction Kit (ProtonDx Ltd, UK), following the manufacturer’s protocol for the extraction of viral DNA/RNA from a liquid sample (Figure 1) (18). Up to 12 extractions at a time (per user) were performed manually using the SmartLid Rack and Vortex Tool (ProtonDx Ltd, UK). See the supplemental section for SmartLid Viral Kit IFU (Supplemental Method 1). The SmartLid extraction protocol required approximately 5–10 minutes per run, whereas the comparator automated platform processed up to 48 samples per run in approximately 15–25 minutes (12,19).

**Figure 1.**
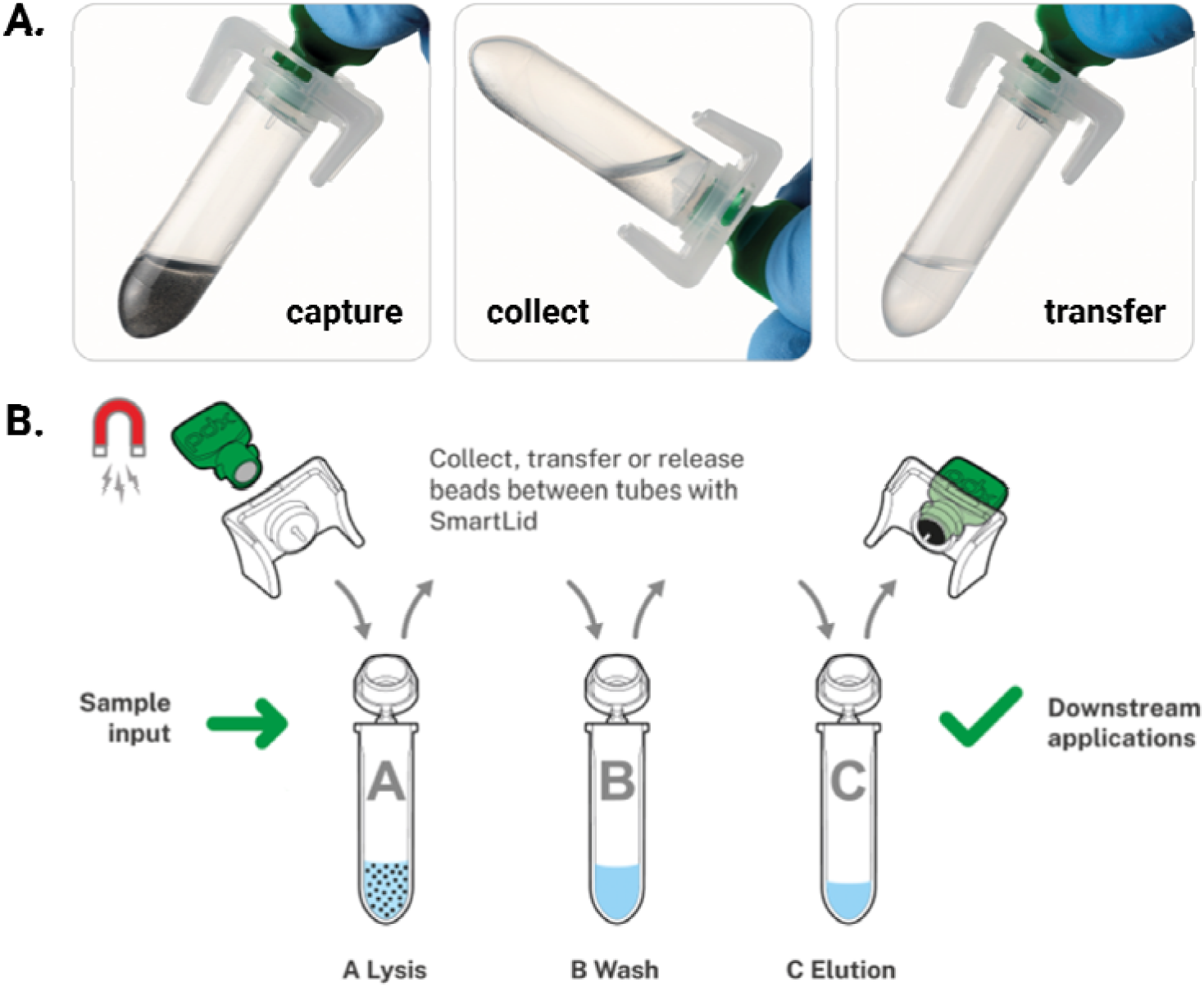
SmartLid magnetic bead handling. (A) Beads are captured, collected, and transferred using a magnet. (B) Beads are shuttled between tubes through lysis, wash, and elution for downstream applications. Created wit BioRender.com

### Downstream amplification and subtyping

Eluates from both extraction methods were tested using separate target-specific, one-step monoplex TaqMan RT-qPCR reactions for influenza A(H1N1)pdm09, influenza A(H3), and influenza B/Victoria. Influenza A subtyping was performed using the CDC Influenza Virus Real-Time RT-PCR Panel for research use only, Influenza A(H3/H1pdm09) Subtyping Panel, version 4 (catalogue FluRUO-16; International Reagent Resource reference FR- 1890; manufactured by the CDC Influenza Division, Atlanta, GA, USA).

Reactions were prepared using the SuperScript™ III Platinum™ One-Step qRT-PCR Kit with ROX (Invitrogen, catalogue 11745500) and amplified on a LightCycler® 480 II instrument (Roche). For each paired specimen, assay reagents, reaction volumes, thermal cycling conditions, fluorescence acquisition settings, and result-interpretation criteria were held constant between extraction methods. Cycle-threshold values were recorded where amplification occurred. Full reaction compositions and thermal cycling conditions are provided in Supplemental Method 2.

### Data analysis

All analyses were conducted in R (version 4.4.1). Sensitivity and specificity were computed for each extraction method in comparison to the study comparator methodology, using two-sided exact binomial 95% confidence intervals (Clopper-Pearson method). The differences in subtyping success among reference-positive specimens were evaluated using an exact McNemar analysis. The Ct analyses were limited to complete paired positive specimens and compared utilising the two-sided Wilcoxon signed-rank test. Rank-order concordance was evaluated by Spearman’s correlation. Subgroup analyses based on influenza subtype and specimen condition employed paired Wilcoxon signed-rank tests with Benjamini-Hochberg adjustment. SmartLid Ct values in specimens identified solely after SmartLid extraction were analysed against those identified by both procedures using the two-sided Wilcoxon rank-sum test. All tests were two-sided, and p < 0.05 was classified as statistically significant.

### Ethical considerations

This study was approved by the Ethics Committee of the East Health Area of Valladolid (Spain) under the code PI-23-3208 in 2023, and follows the terms of the Declaration of Helsinki on the experiments and evaluation of human health data.

## RESULTS

### Qualitative agreement on negative specimens

Both extraction methods yielded complete qualitative agreement on the 47 negative specimens (true negatives 47/47; false positives 0/47), corresponding to an observed specificity of 100% (95% CI: 92.45-100.00) (Table 1).

**Table 1.** Performance comparison of SmartLid and the comparator extraction method on 311 clinical respiratory samples, showing detection outcomes and average Ct values across positive cases.

| Metric | SmartLid | Nextractor® NX-48S |
| --- | --- | --- |
| Reference positives, n | 264 | 264 |
| Reference negatives, n | 47 | 47 |
| Successful subtyping among reference positives | 263 | 241 |
| Unsuccessful subtyping among reference positives | 1 | 23 |
| True negative among reference negatives | 47 | 47 |
| False positives among reference negatives | 0 | 0 |
| Sensitivity (%) [95% CI] | 99.62 [97.91 - 99.99] | 91.29 [87.21 - 94.40] |
| Specificity (%) [95% CI] | 100.00 [92.45 - 100.00] | 100.00 [92.45 - 100.00] |
| Mean Ct [StDev] | 29.33 [4.27] | 31.17 [3.43] |
| Ct range | 20.34 - 38.93 | 23.01 - 39.27 |

### Subtyping success among influenza-positive specimens

Subtyping was successful in 263/264 influenza-positive specimens following SmartLid extraction, for a sensitivity of 99.62% (95% CI: 97.91-99.99). In contrast, subtyping succeeded in 241/264 specimens following extraction by the comparator workflow, for a sensitivity of 91.29% (95% CI: 87.21-94.40). The absolute difference in subtyping success was 8.33 percentage points in favour of SmartLid (Table 1).

A paired contingency analysis clarified the source of this difference: in 23/264 cases (8.71%), subtyping succeeded following SmartLid extraction but failed following the comparator workflow; in 1/264 case (0.38%), the converse was observed; and in 240/264 cases (90.9%), both methods succeeded. No positive specimen failed subtyping with both extraction methods. An exact McNemar test gave P = 2.98 × 10⁻⁶ (continuity-corrected χ² = 18.38, P = 1.8 × 10⁻⁵); the absolute difference in subtyping success was 8.33 percentage points (95% CI 4.84 to 11.83).

Taken together, these results indicate that subtyping failure was substantially less frequent after SmartLid extraction (1/264, 0.38%) than after the comparator workflow (23/264, 8.71%) under otherwise identical amplification conditions, consistent with improved analytical sensitivity at or near assay detection limits.

### Quantitative Ct performance in paired specimens

Ct analysis was restricted to the 240 influenza-positive specimens with complete paired measurements from both extraction methods. Ct values were consistently lower following SmartLid extraction than following the comparator workflow. The median paired difference in Ct (SmartLid minus comparator) was −2.78 cycles, indicating greater accessible nucleic-acid yield after SmartLid extraction. A paired Wilcoxon signed-rank test demonstrated a highly significant shift in Ct values in favour of SmartLid (p < 0.001; estimated paired location shift −2.60 cycles, 95% CI −2.82 to −2.37) (Figure 2A).

**Figure 2.**
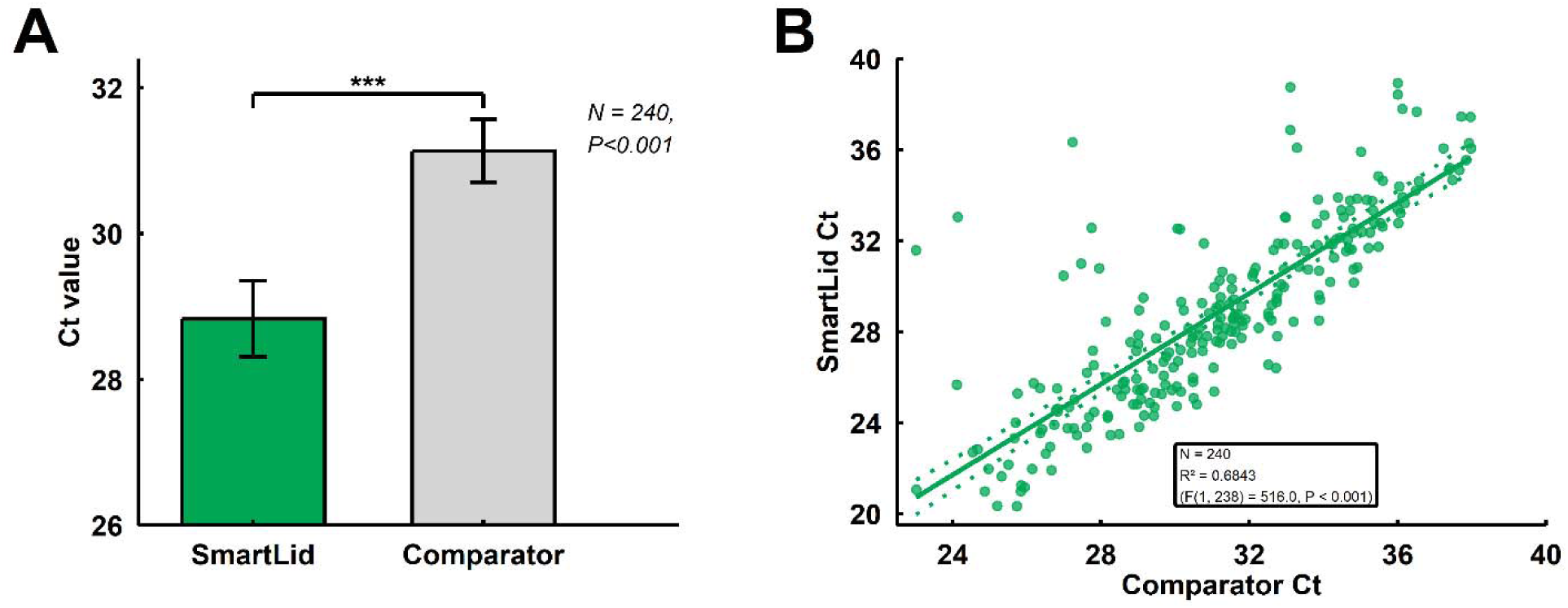
Performance comparison of SmartLid and comparator extraction methods across influenza-positive clinical specimens. (A) Comparison of Ct values obtained from SmartLid and the comparator extraction across complete paired influenza-positive clinical samples (n = 240). Bars represent mean +/- 95% CI. Statistical significance was assessed using a paired Wilcoxon signed-rank test (*** p < 0.001). (B) Concordance of Ct values between SmartLid and the comparator method in complete-case paired samples (n = 240). The solid line is the ordinary least-squares regression of SmartLid on comparator Ct, with its 95% confidence bounds shown as dotted lines (R² = 0.6843; F(1, 238) = 516.0; P < 0.001). Rank ordering of specimens was preserved between methods (Spearman rho = 0.84, P < 0.001).

Rank-order concordance between methods was strong (Spearman rho = 0.84, p < 0.001), indicating that, while SmartLid shifted Ct values downward overall, relative sample-to-sample signal ordering was preserved (Figure 2B).

### Subgroup analyses by influenza subtype and specimen condition

The Ct advantage of SmartLid was preserved across viral subtypes and storage conditions. In analyses stratified by subtype-B/Victoria (BVic; n = 100), A/H3 (H3; n = 51), and A/H1 (H1; n = 89)-paired Ct values were significantly lower with SmartLid than with the comparator workflow in each subgroup after Benjamini-Hochberg correction (all adjusted p < 0.001, paired Wilcoxon tests). Likewise, when stratified by specimen condition, both fresh (n = 148) and frozen (n = 92) specimens showed significantly lower paired Ct values after SmartLid extraction (both adjusted p < 0.001). Effect directions were consistent across all strata, and no stratum showed attenuation of the Ct advantage (Figure 3).

**Figure 3.**
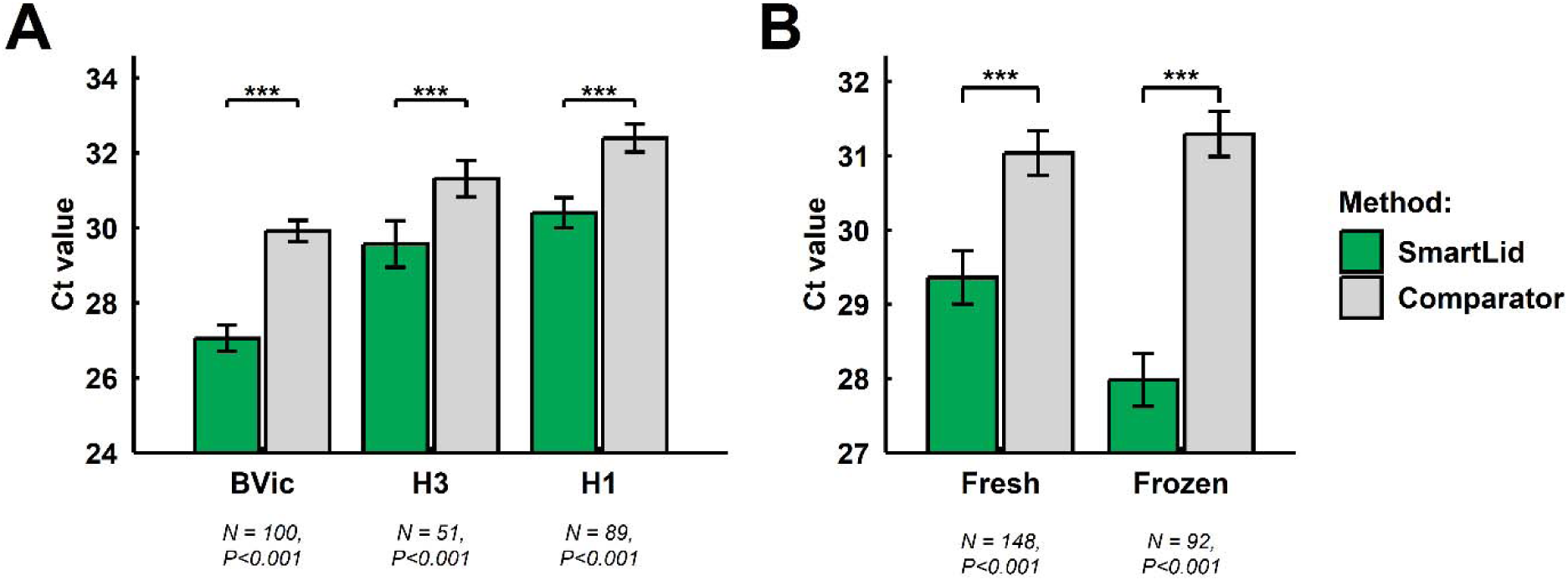
Comparison of Ct values obtained using the SmartLid and comparator extraction methods, stratified by influenza subtype and specimen condition. (A) Paired Ct values by subtype: BVic, B/Victoria; H3, influenza A(H3); H1, influenza A(H1N1)pdm09. (B) Paired Ct values by specimen condition (fresh versus frozen). Analyses were restricted, within each stratum, to specimens returning a valid Ct from both extraction methods; specimens lacking a matched value were excluded to preserve pairing (A: n = 100, 51 and 89; B: n = 148 and 92). Bars represent mean ± SEM. Statistical significance was assessed using paired Wilcoxon signed-rank tests with Benjamini–Hochberg correction applied across all five strata (*** adjusted P < 0.001). Figures were generated in R version 4.4.1 using ggplot2 and patchwork.

### Improved subtyping of low viral load samples with SmartLid

Samples that were negative with the comparator extraction but positive with SmartLid showed substantially higher Ct values than specimens successfully detected by both methods (median Ct 34.37 versus 28.54; Wilcoxon rank-sum p < 0.001; n = 23 vs 240). This pattern is consistent with SmartLid extraction exhibiting greater analytical sensitivity than the comparator protocol, particularly near the assay detection limit.

## DISCUSSION

This head-to-head study shows that SmartLid matches the specificity of the Nextractor^®^ NX-48S automated platform while delivering a clear gain in analytical sensitivity for influenza subtyping under routine public-health laboratory conditions. Across 311 clinical specimens run with identical downstream assays, SmartLid maintained 100% specificity and achieved significantly higher subtyping success on positives than the automated comparator. Quantitatively, SmartLid yielded significantly lower Ct values with rank-ordering of specimens preserved between methods and the advantage present in every subtype and storage stratum examined.

Together, these results indicate that extraction efficiency was the primary driver of the performance difference and that SmartLid increases the likelihood of successful subtyping, especially near the assay’s detection limit (20).

### Analytical performance and clinical relevance

The 8.33-percentage-point absolute increase in subtyping success is operationally meaningful. Most paired positives were concordant (240/264), implying that both workflows perform similarly at moderate-to-high template abundance. The imbalance among discordant pairs and the observation that comparator- negative/SmartLid-positive specimens had substantially higher SmartLid Cts than dual-detected samples (median 34.37 vs. 28.54; p < 0.001) pinpoint the low-template regime as the setting where extraction efficiency is outcome-defining (21,22). Recovering these borderline detections increases subtyping completeness for surveillance, reduces the need for repeat sampling or reflex testing, and may improve timeliness of public-health decision-making during periods of heightened activity.

Importantly, sensitivity gains were not accompanied by loss of specificity: both methods produced zero false positives among 47 negatives. This supports that lower Cts after SmartLid reflect genuine increases in accessible template rather than relaxed calling thresholds or contamination (23).

### Operational implications and use-case segmentation

SmartLid’s simple, manual design enables deployment beyond central laboratories, supporting surveillance in resource-limited settings with minimal training and no reliance on centrifugation or complex infrastructure. The observed analytical gains strengthen the case for its use in decentralised testing, surge capacity, or contexts with staffing and maintenance constraints (12–14). By contrast, the automated comparator method remains advantageous for large centres such as NICV, where high-throughput, walk-away batch processing, LIMS integration, and standardised, auditable workflows are priorities (15). In practice, the approaches are complementary: SmartLid could expand reach and rescue low-load detection at the edge of assay sensitivity, while automated platforms provide efficient routine processing at scale. A hub-and-spoke model, implementing SmartLid for field or satellite intake and automated systems for central confirmatory or high-volume work, could maximise coverage and resilience (10).

A key benefit of deploying SmartLid at or near the POC is the dramatic reduction in time to diagnosis and treatment for patients. When testing is done on-site with rapid methods, clinicians can make decisions immediately. For example, a recent real-world study in U.S. clinics compared on-site molecular testing to conventional laboratory testing for respiratory infections: POC testing yielded a median time to influenza diagnosis of 0 days versus 5 days with lab-based testing (24). This accelerated timeline can be lifesaving in many cases; identifying an influenza infection on the spot allows antiviral medications to be started during the critical early window, improving patient recovery and reducing complications (24). Faster diagnosis can also mean quicker implementation of isolation precautions or contact tracing, limiting spread in hospitals and communities. Consistent with this, randomised and real-world evaluations of molecular point-of-care testing show earlier use of isolation facilities and ∼42% fewer droplet-precaution days, alongside shorter overall hospital stays (25–27).

Improved subtyping success and faster time-to-result could also have direct therapeutic advantages. When influenza B is identified, baloxavir may be preferred over oseltamivir, supported by CAPSTONE-2 subgroup findings of faster clinical improvement in high-risk outpatients and evidence of lower hospitalisation and healthcare utilisation (28). Single-dose administration also aids adherence, making a rapid “B” call clinically actionable (28,29). During documented H275Y A(H1N1)pdm09 variant circulation, identifying A(H1N1)pdm09 may flag possible oseltamivir resistance and support consideration of zanamivir or baloxavir, which retain activity (30,31). In this context, SmartLid’s higher subtyping yield, especially near assay limits, can convert otherwise indeterminate cases into subtype-directed prescribing decisions at the point of care.

SmartLid strengthens influenza surveillance by enabling accurate, manual extraction at or near the POC, bringing reliable subtyping to clinics and field posts and closing geographic and sensitivity gaps. By delivering rapid, field-ready extraction and higher-quality PCR input, it could accelerate signal detection (local spikes, drifted/novel strains) and feed richer data into the WHO GISRS pipeline that underpins vaccine-strain selection. Operationally, decentralising extraction adds surge capacity in a hub-and-spoke model while preserving specificity, improving situational awareness without overloading central labs (24,32–34).

### Limitations and future directions

This study was conducted at a single centre using one automated comparator and a subtyping panel limited to A(H1N1)pdm09, A(H3), and B/Victoria. Further multicentre evaluations should therefore assess performance across additional extraction systems, instruments, specimen types, and circulating influenza strains (35).

Optimising SmartLid for automated and semi-automated platforms should be the main priority to be explored. This would bridge manual-to-automated workflows, enabling larger centres to realise SmartLid’s sensitivity gains in high-throughput use without disrupting existing robotics/workflows (36,37).

In parallel, SmartLid chemistry could be adapted for compatibility with established high-throughput magnetic-particle processors, particularly the KingFisher Flex. Given the platform’s widespread use in high-throughput SARS-CoV-2 testing during the COVID-19 pandemic, such compatibility could facilitate adoption by laboratories with existing instrumentation and workflows while preserving the improved Ct performance observed with SmartLid (36,38).

## Conclusions

Under routine influenza surveillance conditions, SmartLid improved subtyping success and lowered Ct values relative to an established automated extractor without compromising specificity. Its decentralised, rapid operation provides a practical route to extend high-quality molecular subtyping beyond central laboratories, while automated high-throughput systems remain desirable for large-scale processing. Leveraging both modalities can enhance surveillance coverage, responsiveness, and equity of access to molecular diagnostics.

## Data Availability

All data produced in the present study are available upon reasonable request to the authors

## Acknowledgements

This work was supported by the Jameel Fund for Infectious Disease Research and Innovation, and the CAMO-Net programme funded by the Wellcome Trust (grant number 226691/Z/22/Z). We also acknowledge the support of the National Institute for Health Research (NIHR) Biomedical Research Centre at Imperial College London, and the Centre for Antimicrobial Optimisation at Imperial College London funded by the UK Department of Health and Social Care.

## Author Contributions

KSL, JSEB, JRM, and ISM contributed to the conception and design of the study. MLC, SSP, JRM, and ISM contributed to the initial drafting of the manuscript, interpreting the results, and accessed, verified, and reviewed the raw data. JSM, CRC, IP and KMC contributed to data acquisition and analysis. AHH and JRM contributed to securing funding. All authors contributed to data interpretation and critically revised the manuscript for important intellectual content. All authors had full access to all study data and accept final responsibility for the decision to submit the manuscript for publication.

## REFERENCES

1. Gharpure R, Chard AN, Cabrera Escobar M, Zhou W, Valleau MM, Yau TS, et al. Costs and cost-effectiveness of influenza illness and vaccination in low- and middle-income countries: A systematic review from 2012 to 2022. PLoS Med. 2024 Jan 5;21(1):e1004333. doi:10.1371/journal.pmed.1004333

2. Iuliano AD, Roguski KM, Chang HH, Muscatello DJ, Palekar R, Tempia S, et al. Estimates of global seasonal influenza-associated respiratory mortality: a modelling study. The Lancet. 2018 Mar;391(10127):1285–300. doi:10.1016/S0140-6736(17)33293-2

3. Ramos-Rincón JM, Pinargote-Celorio H, González-de-la-Aleja P, Sánchez-Payá J, Reus S, Rodríguez-Díaz JC, et al. Impact of influenza related hospitalization in Spain: characteristics and risk factor of mortality during five influenza seasons (2016 to 2021). Front Public Health. 2024 Apr 2;12:1360372. doi:10.3389/fpubh.2024.1360372

4. Sanz-Muñoz I, Arroyo-Hernantes I, Martín-Toribio A, Toquero-Asensio M, Sánchez-Martínez J, Rodríguez-Crespo C, et al. Disease burden of influenza in Spain: A five- season study (2015–2020). Hum Vaccin Immunother. 2025 Dec 31;21(1). doi:10.1080/21645515.2024.2440206

5. Maltezou HC, Sourri F, Lemonakis N, Karapanou A, Giannouchos T V, Gamaletsou MN, et al. Evaluation of the influenza-like illness case definition and the acute respiratory infection case definition in the diagnosis of influenza and COVID-19 in healthcare personnel. Infect Dis Health. 2025 Feb 1;30(1):23–7. doi:10.1016/j.idh.2024.08.002

6. World Health Organization. Implementing the integrated sentinel surveillance of influenza and other respiratory viruses of epidemic and pandemic potential by the Global Influenza Surveillance and Response System. Geneva: World Health Organization; 2024. 66 p. ISBN 978-92-4-010143-2. Licence: CC BY-NC-SA 3.0 IGO. Available from: https://www.who.int/publications/i/item/9789240101432

7. Qian Q, Fan G, Yang W, Shen C, Yang Y, Liu Y, et al. Advances in Diagnostic Techniques for Influenza Virus Infection: A Comprehensive Review. Trop Med Infect Dis. 2025 May 28;10(6). doi:10.3390/tropicalmed10060152

8. Chaves M, Hashish A, Osemeke O, Sato Y, Suarez DL, El-Gazzar M. Evaluation of Commercial RNA Extraction Protocols for Avian Influenza Virus Using Nanopore Metagenomic Sequencing. Viruses. 2024 Sep 7;16(9):1429. doi:10.3390/v16091429

9. Yu F, Qiu T, Zeng Y, Wang Y, Zheng S, Chen X, et al. Comparative Evaluation of Three Preprocessing Methods for Extraction and Detection of Influenza A Virus Nucleic Acids from Sputum. Front Med (Lausanne). 2018 Mar 2;5:56. doi:10.3389/fmed.2018.00056

10. Centers for Disease Control and Prevention. CDC Health Alert Network Advisory No. CDCHAN-00520 [Internet]. 2025 [cited 2026 May 5]. Accelerated Subtyping of Influenza A in Hospitalized Patients. Available from: https://www.cdc.gov/han/2025/han00520.html

11. Zhang YB, Arizti-Sanz J, Bradley A, Huang Y, Kosoko-Thoroddsen TSF, Sabeti PC, et al. CRISPR-Based Assays for Point-of-Need Detection and Subtyping of Influenza. J Mol Diagn. 2024 Jul;26(7):599–612. doi:10.1016/j.jmoldx.2024.04.004

12. Pennisi I, Cavuto ML, Miglietta L, Malpartida-Cardenas K, Stringer OW, Mantikas KT, et al. Rapid, Portable, and Electricity-free Sample Extraction Method for Enhanced Molecular Diagnostics in Resource-Limited Settings. Anal Chem. 2024 Jul 16;96(28):11181–8. doi:10.1021/acs.analchem.4c00319

13. Cavuto ML, Malpartida-Cardenas K, Pennisi I, Pond MJ, Mirza S, Moser N, et al. Portable molecular diagnostic platform for rapid point-of-care detection of mpox and other diseases. Nat Commun. 2025 Mar 24;16(1):2875. doi:10.1038/s41467-025-57647-3

14. Rakotomalala Robinson D, Pennisi I, Cavuto ML, Kiemde F, Chamai M, Some DY, et al. Sensitive near point-of-care detection of asymptomatic and submicroscopic Plasmodium falciparum infections in African endemic countries. Nat Commun. 2025 Oct 10;16(1):8925. doi:10.1038/s41467-025-64027-4

15. Sanz-Muñoz I, Sánchez-Martínez J, Rodríguez-Crespo C, Arroyo-Hernantes I, Domínguez-Gil M, Rojo-Rello S, et al. Association of viral loads of influenza A (H3N2) with age and care setting on presentation—a prospective study during the 2022-2023 influenza season in Spain. International Journal of Infectious Diseases. 2024 Jun;143:107034. doi:10.1016/j.ijid.2024.107034

16. Deiana M, Mori A, Piubelli C, Scarso S, Favarato M, Pomari E. Assessment of the direct quantitation of SARS-CoV-2 by droplet digital PCR. Sci Rep. 2020 Oct 30;10(1):18764. doi:10.1038/s41598-020-75958-x

17. Leber AL, Everhart K, Daly JA, Hopper A, Harrington A, Schreckenberger P, et al. Multicenter Evaluation of BioFire FilmArray Respiratory Panel 2 for Detection of Viruses and Bacteria in Nasopharyngeal Swab Samples. J Clin Microbiol. 2018 Jun;56(6). doi:10.1128/JCM.01945-17

18. ProtonDx. Viral DNA/RNA Extraction Starter Kit: Resources and Training Videos [Internet]. [cited 2026 Apr 7]. Available from: https://www.protondx.com/resources-smartlid

19. Genolution. Nextractor NX-48S [Internet]. Republic of Korea: Genolution; [cited 2026 Apr 7]. Available from: https://genolution.co.kr/nextractor-nx-48s

20. Kim S, Lee W. Does McNemar’s test compare the sensitivities and specificities of two diagnostic tests? Stat Methods Med Res. 2017 Feb;26(1):142–54. doi:10.1177/0962280214541852

21. Komiazyk M, Walory J, Kozinska A, Wasko I, Baraniak A. Impact of the Nucleic Acid Extraction Method and the RT-qPCR Assay on SARS-CoV-2 Detection in Low-Viral Samples. Diagnostics. 2021 Nov 30;11(12):2247. doi:10.3390/diagnostics11122247

22. Gdoura M, Abouda I, Mrad M, Ben Dhifallah I, Belaiba Z, Fares W, et al. SARS-CoV2 RT-PCR assays: In vitro comparison of 4 WHO approved protocols on clinical specimens and its implications for real laboratory practice through variant emergence. Virol J. 2022 Dec 28;19(1):54. doi:10.1186/s12985-022-01784-4

23. Alonzo TA, Brinton JT, Ringham BM, Glueck DH. Bias in estimating accuracy of a binary screening test with differential disease verification. Stat Med. 2011 Jul 10;30(15):1852–64. doi:10.1002/sim.4232

24. Stockl KM, Tucker J, Beaubrun A, Certa JM, Becker L, Chase JG. Real-world use of multiplex point-of-care molecular testing or laboratory-based molecular testing for influenza-like illness in a 2021 to 2022 US outpatient sample. PLoS One. 2024 Nov 11;19(11):e0313660. doi:10.1371/journal.pone.0313660

25. Muller MP, Junaid S, Matukas LM. Reduction in total patient isolation days with a change in influenza testing methodology. Am J Infect Control. 2016 Nov;44(11):1346–9. doi:10.1016/j.ajic.2016.03.019

26. Clark TW, Beard KR, Brendish NJ, Malachira AK, Mills S, Chan C, et al. Clinical impact of a routine, molecular, point-of-care, test-and-treat strategy for influenza in adults admitted to hospital (FluPOC): a multicentre, open-label, randomised controlled trial. Lancet Respir Med. 2021 Apr;9(4):419–29. doi:10.1016/S2213-2600(20)30469-0

27. Berry L, Lansbury L, Gale L, Carroll AM, Lim WS. Point of care testing of Influenza A/B and RSV in an adult respiratory assessment unit is associated with improvement in isolation practices and reduction in hospital length of stay. J Med Microbiol. 2020 May 1;69(5):697–704. doi:10.1099/jmm.0.001187

28. Ison MG, Portsmouth S, Yoshida Y, Shishido T, Mitchener M, Tsuchiya K, et al. Early treatment with baloxavir marboxil in high-risk adolescent and adult outpatients with uncomplicated influenza (CAPSTONE-2): a randomised, placebo-controlled, phase 3 trial. Lancet Infect Dis. 2020 Oct;20(10):1204–14. doi:10.1016/S1473-3099(20)30004-9

29. Ishiguro N, Morioka I, Nakano T, Manabe A, Kawaguchi K, Tanaka S, et al. Clinical and Virologic Outcomes of Baloxavir Compared with Oseltamivir in Pediatric Patients with Influenza in Japan. Infect Dis Ther. 2025 Apr 28;14(4):833–46. doi:10.1007/s40121-025-01131-4

30. Takashita E, Shimizu K, Usuku S, Senda R, Okubo I, Morita H, et al. An outbreak of influenza A(H1N1)pdm09 antigenic variants exhibiting cross-resistance to oseltamivir and peramivir in an elementary school in Japan, September 2024. Eurosurveillance. 2024 Dec 12;29(50). doi:10.2807/1560-7917.ES.2024.29.50.2400786

31. Hussain S, Meijer A, Govorkova EA, Dapat C, Gubareva L V., Barr IG, et al. Global update on the susceptibilities of influenza viruses to neuraminidase inhibitors and the cap-dependent endonuclease inhibitor baloxavir, 2020–2023. Antiviral Res. 2025 Sep;241:106217. doi:10.1016/j.antiviral.2025.106217

32. Hay AJ, McCauley JW. The WHO global influenza surveillance and response system (GISRS)—A future perspective. Influenza Other Respir Viruses. 2018 Sep 25;12(5):551–7. doi:10.1111/irv.12565

33. Shu Y, McCauley J. GISAID: Global initiative on sharing all influenza data – from vision to reality. Eurosurveillance. 2017 Mar 30;22(13). doi:10.2807/1560-7917.ES.2017.22.13.30494

34. Gupta S, Gupta T, Gupta N. Global respiratory virus surveillance: strengths, gaps, and way forward. International Journal of Infectious Diseases. 2022 Aug;121:184–9. doi:10.1016/j.ijid.2022.05.032

35. Lane K, Palm ME, Marion E, Kay MT, Thompson D, Stroud M, et al. Approaches for enhancing the informativeness and quality of clinical trials: Innovations and principles for implementing multicenter trials from the Trial Innovation Network. J Clin Transl Sci. 2023 May 25;7(1):e131. doi:10.1017/cts.2023.560

36. Lim HJ, Jung HS, Park MY, Baek YH, Kannappan B, Park JY, et al. Evaluation of Three Automated Extraction Systems for the Detection of SARS-CoV-2 from Clinical Respiratory Specimens. Life (Basel). 2022 Jan 4;12(1):68. doi:10.3390/life12010068

37. Fang X, Willis RC, Burrell A, Evans K, Hoang Q, Xu W, et al. Automation of Nucleic Acid Isolation on KingFisher Magnetic Particle Processors. JALA: Journal of the Association for Laboratory Automation. 2007 Aug 1;12(4):195–201. doi:10.1016/j.jala.2007.05.001

38. Thom RE, Eastaugh LS, O’Brien LM, Ulaeto DO, Findlay JS, Smither SJ, et al. Evaluation of the SARS-CoV-2 Inactivation Efficacy Associated With Buffers From Three Kits Used on High-Throughput RNA Extraction Platforms. Front Cell Infect Microbiol. 2021 Sep 16;11. doi:10.3389/fcimb.2021.716436

