## Supplemental Methods for "Rapid magnetic bead nucleic acid extraction enhances influenza RT-qPCR sensitivity and subtyping success"

‡ These authors contributed equally

### SmartLid Extraction Protocol

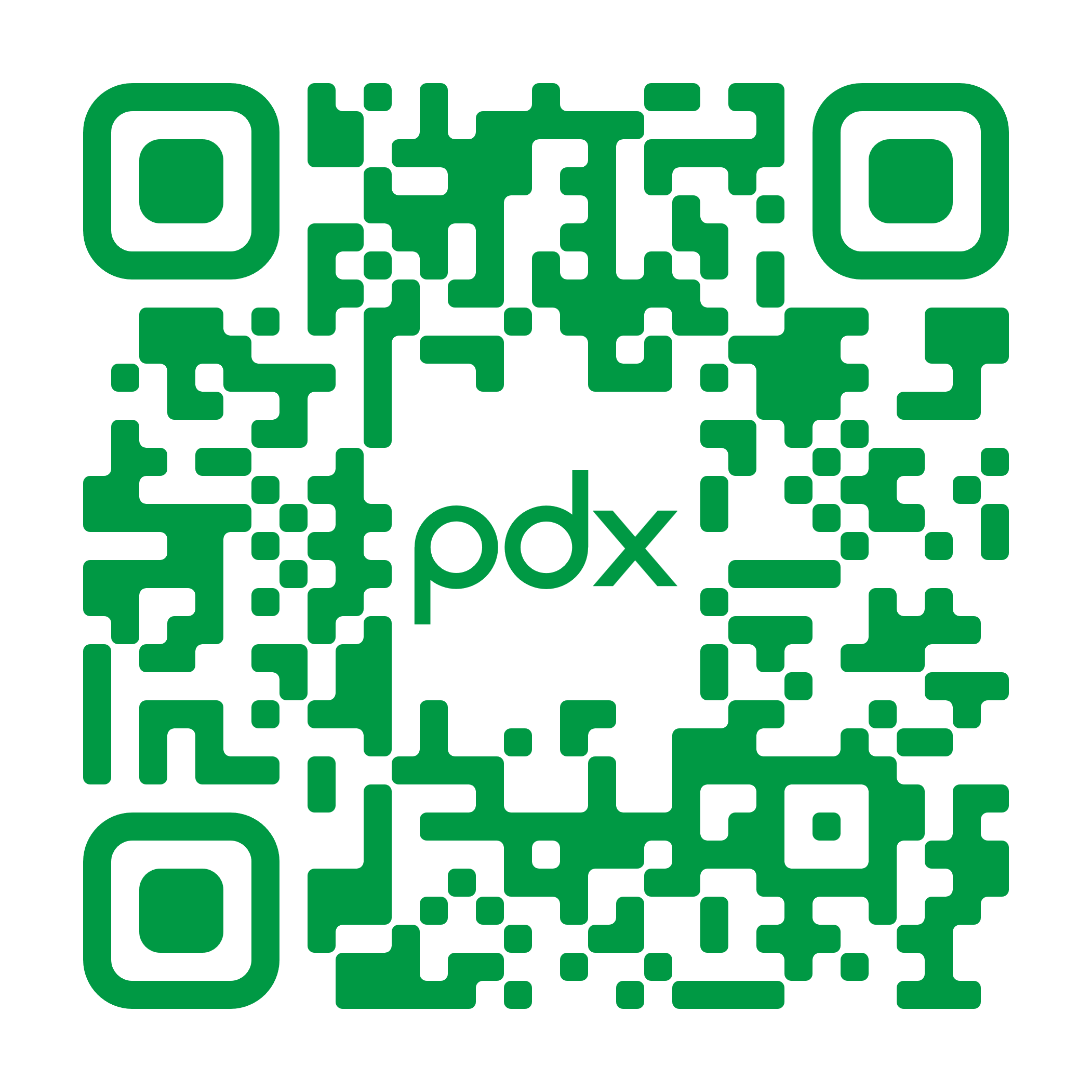

**Resources and Training Videos**

**IMPORTANT Before Starting**

When opening a new SmartLid Viral DNA/RNA Extraction Kit, **add 24 mL of molecular biology grade ethanol (>99%) to the Wash Buffer (concentrate)** as indicated on the bottle. There is a box on the bottle label to check once this is completed.

*Always ensure all buffers are liquid and homogeneous prior to use. In the case of visible precipitate, redissolve by warming and gentle mixing.*

**If using the SmartLid Viral DNA/RNA Extraction Kit for the first time, we recommend watching our instructional videos by visiting** [**www.protondx.com**](http://www.protondx.com) **or scanning the QR code above.**

**SETUP for Multiple Extractions**

1. Set up the required number of flip-cap tubes in a rack (Recommended: SmartLid Rack 100175), with different rows for **Lysis** (Row A), **Wash** (Row B), and **Elution** (Row C). For example, 6 extractions will require a total of 18 tubes, split in 3 rows of 6 tubes.
2. If extracting from a **liquid sample**, we recommend creating a master mix of Lysis Binding Buffer and Magnetic Beads to ensure more consistent extractions. Prepare this master mix according to ratios in the table below, and multiply by the number of extractions:

| **Component** | **Volume per tube ^[1]^** | **Volume of Master Mix for n extractions** |
| --- | --- | --- |
| Lysis Binding Buffer | 700 µL | 700 µL *×* **n** |
| Magnetic Beads | 20 µL ^[2]^ | 20 µL *×* **n** ^[2]^ |
| **Total volume** | 720 µL ^[2]^ | 720 µL *×* **n** ^[2]^ |

^[1] Use 10% overage calculation when making a master mix for use with multiple samples.^

^[2] Up to 40 µL of Magnetic Beads per sample can be used to improve yield if necessary, resulting in a master mix volume per extraction of 740 µL.^

**Note:** If extracting directly from dry swabs without a liquid transport medium, skip this step (step 2) and follow the second half of the **Lysis Binding** section below (“Extracting from dry swabs without transport medium”).

3. Prefill all tubes according to the table below:

| **Tube/Step** | **Components and Volumes** | |
| --- | --- | --- |
| A: Lysis Binding | ***From Liquid Media:***  **720 µL** ^[1]^ Lysis Binding Master Mix | ***From Dry Swab:***  **700 µL** Lysis Binding Buffer **ONLY***  **(*Magnetic Beads will be added later.)** |
| B: Wash | **300 µL** Wash Buffer ^[2]^ | |
| C: Elution | **50 µL** ^[3]^ Elution Buffer | |

^[1] See note above regarding Magnetic Bead concentration/volume.^

^[2] Ensure that 24 mL of molecular biology grade ethanol (>99%) has been added to the Wash Buffer (concentrate) as indicated on the bottle.^

^[3] As little as^ **^30 µL^** ^can be used. While this will result in a more concentrated purified sample, final recovery may be less than 30 µL.^ *^DO NOT centrifuge tubes with SmartLids inserted in order to recover more elution from the lid.^*

**LYSIS BINDING**

**Extraction from liquid sample:**

1. Add up to **200 µL** of sample to the lysis tube (prefilled with Lysis Binding Buffer and Magnetic Beads master mix).
2. Close with a SmartLid (without a Magnetic Key inserted) and briefly shake the tube (**1-2 times**) to ensure the sample and Lysis Binding Buffer is sufficiently mixed.
3. Wait **2-5 minutes** for complete lysis of the sample.

**Note:** For any sample stored and preserved in lysing/inactivating media (e.g., Copan eNat^®^ Buffer or similar), skip this step and proceed directly to Step 4.

4. Temporarily remove the SmartLid and add **500 µL** of IPA (>99%) to the lysed sample.
5. Return the SmartLid (still without the Magnetic Key) to the tube and mix/shake for **60 seconds**.

**Note:** For this mixing/shaking step, and all to follow, the SmartLid Shaker (100173) can be used to conveniently enable simultaneous mixing of up to 12 samples at a time. Always ensure that the retainer arm of the SmartLid Shaker is latched securely before shaking. (See instructional video for more information at [www.protondx.com](http://www.protondx.com) or the QR code above.)

6.
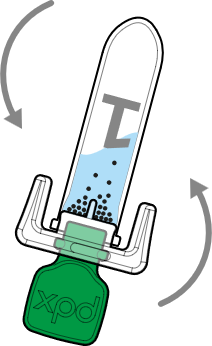
Insert a Magnetic Key into the SmartLid, turning it 90 degrees clockwise to lock, and invert the tubes several times to collect all Magnetic Beads onto the SmartLid.

**Note:** This collection step is the longest of the entire protocol, due to the large volume and high viscosity of the Lysis Binding Buffer. We recommend allowing the tube to remain upside down for **~30 seconds** after the first inversion, followed by multiple quicker (**~2-3 seconds**) inversions until the liquid is clear and all Magnetic Beads are collected.

**Extraction from dry swabs without transport medium:**

1. Carefully place the tip of the swab into the lysis tube (with **700 µL** of Lysis Binding Buffer), swirl for **30 seconds,** then remove.
2. Close with a SmartLid and briefly shake the tube (**1-2 times**) to ensure the sample and Lysis/Binding Buffer is sufficiently mixed.
3. Wait **2-5 minutes** for complete lysis of the sample.
4. Add **500 µL** of IPA (>99%) and **20-40 µL** of Magnetic Beads.
5. Follow steps 5-6 from above to complete the lysis/binding process.

**Protocol continues on the next page 🡪**

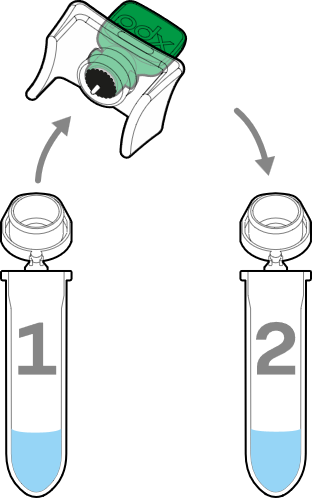

**WASH**

1. Transfer the SmartLid (with Magnetic Key INSERTED) from the lysis tube into the corresponding wash tube, ensuring it’s firmly inserted in the wash tube.

**Note:** If the Magnetic Key is not inserted during transfer of the SmartLid, the Magnetic Beads can fall off, and the captured nucleic acids will be lost.

2. Remove the Magnetic Key and mix/shake the tube for **60 seconds**, ensuring that all Magnetic Beads are fully resuspended.
3. Insert/lock the Magnetic Key into the SmartLid and invert the tube several times to collect all Magnetic Beads onto the SmartLid. Ensure the liquid is clear before proceeding.

**Note 1:** It can help to initially shake the tube with the Magnetic Key inserted to first resuspend all Magnetic Beads, before gently inverting to collect the beads onto the SmartLid.

**Note 2:** There is enough Wash Buffer provided to enable a second washing step. If a second wash is desired, remove the SmartLid (setting it down on any clean surface), discard the old wash buffer appropriately, refill the tube with **300 µL** of wash buffer, and repeat steps 1-3 above.

4. Once all washing steps are complete, and all Magnetic Beads are collected onto the SmartLid, remove the SmartLid and set it down (Magnetic Beads facing down) on a clean surface for **60 seconds** to **allow all EtOH to evaporate.**

**ELUTION**

1. Once the evaporation step is complete, insert the SmartLid (with Magnetic Key still INSERTED) into the corresponding elution tube.
2. Remove the Magnetic Key and mix/shake the tube for **60 seconds**, ensuring that all Magnetic Beads are fully resuspended.

**Note:** Even though the liquid volume is significantly smaller than in previous steps, there is no need to mix/shake any differently, even when working with <50 µL.

3. Insert/lock the Magnetic Key into the SmartLid. This time, due to the small liquid volume for the elution step, after gently inverting 1-2 times, **shake the tube to fully collect all Magnetic Beads.**

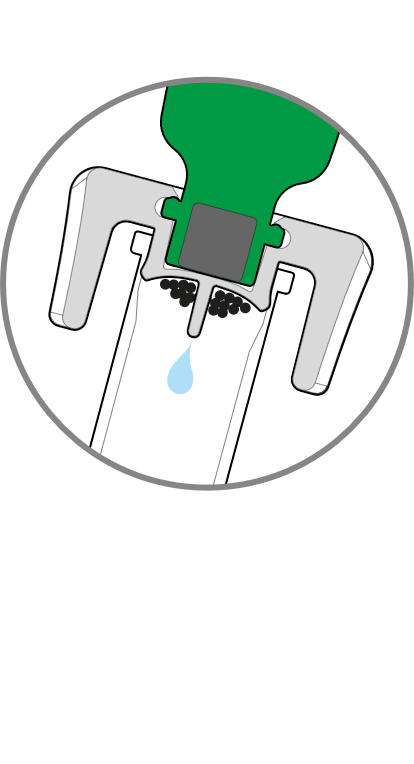

4. Flick down the tube (do not centrifuge) to collect as much elution volume as possible, discard the SmartLid and attached Magnetic Beads (all nucleic acids are now released into solution), and place the closed elution tube in a cold block or on ice until further use.

**Note:** Retain the Magnetic Keys for future extractions. They can be reused indefinitely to limit disposable waste.

**The purified and eluted nucleic acids are now ready for immediate use in downstream applications.**

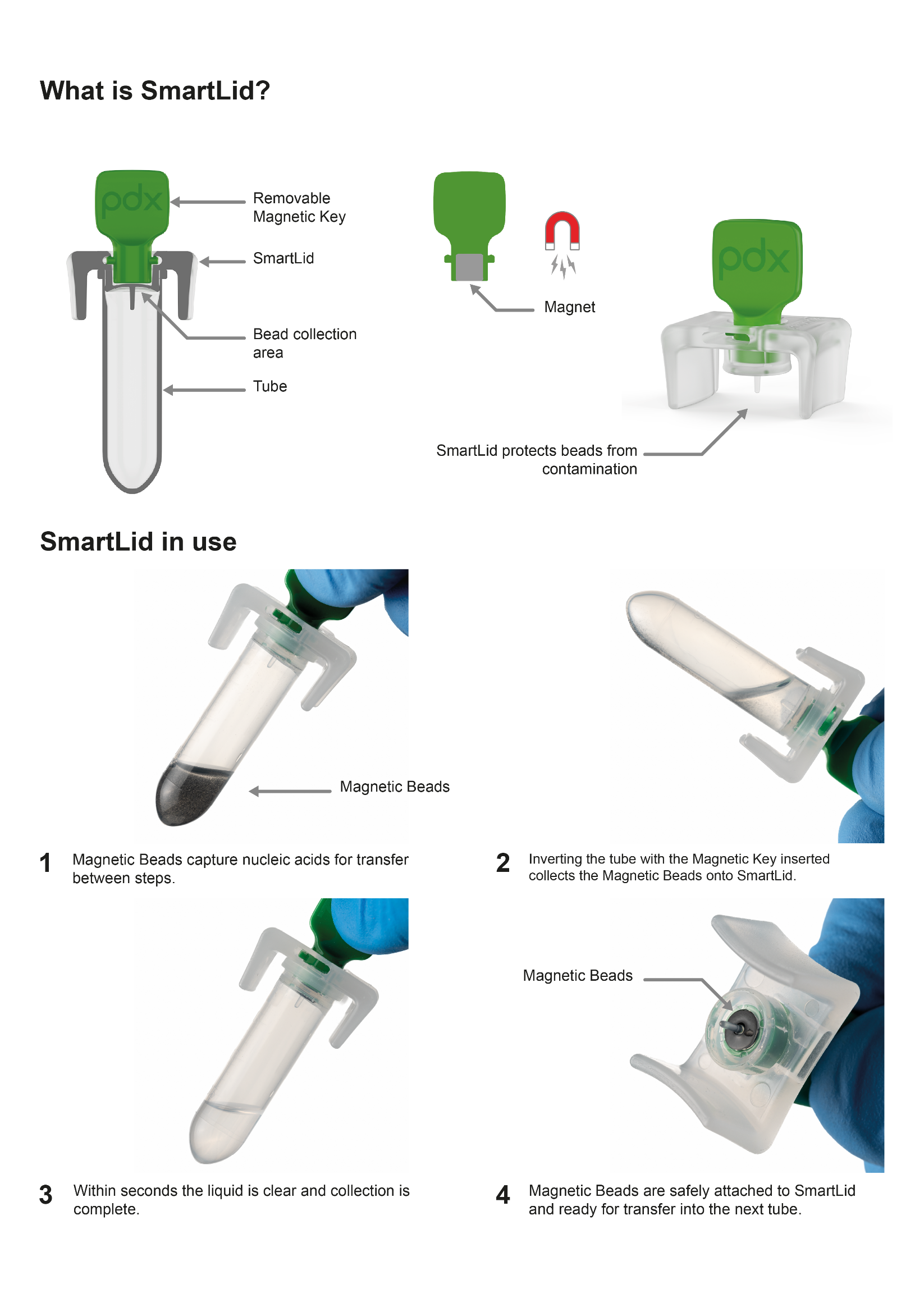

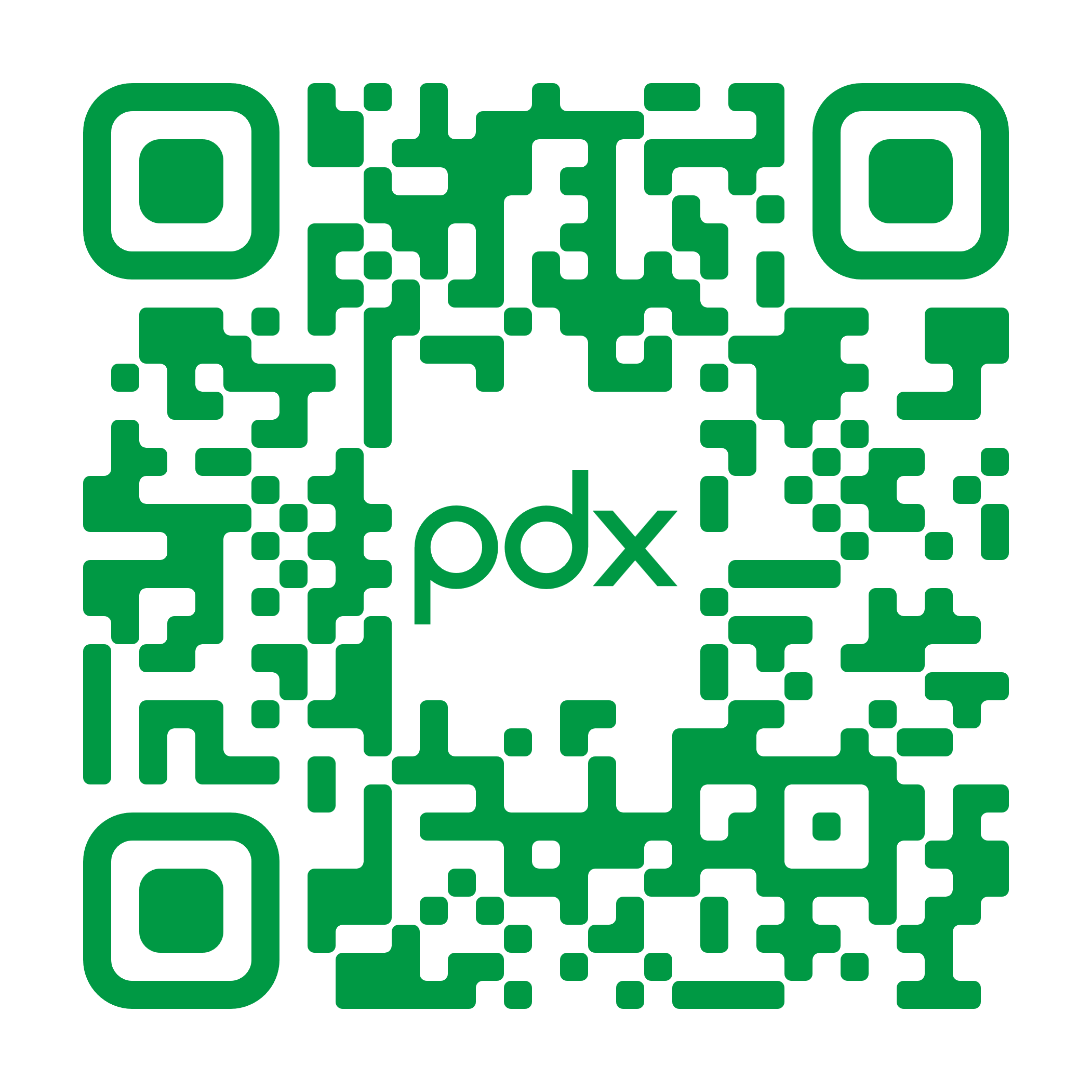

**Resources and Training Videos**

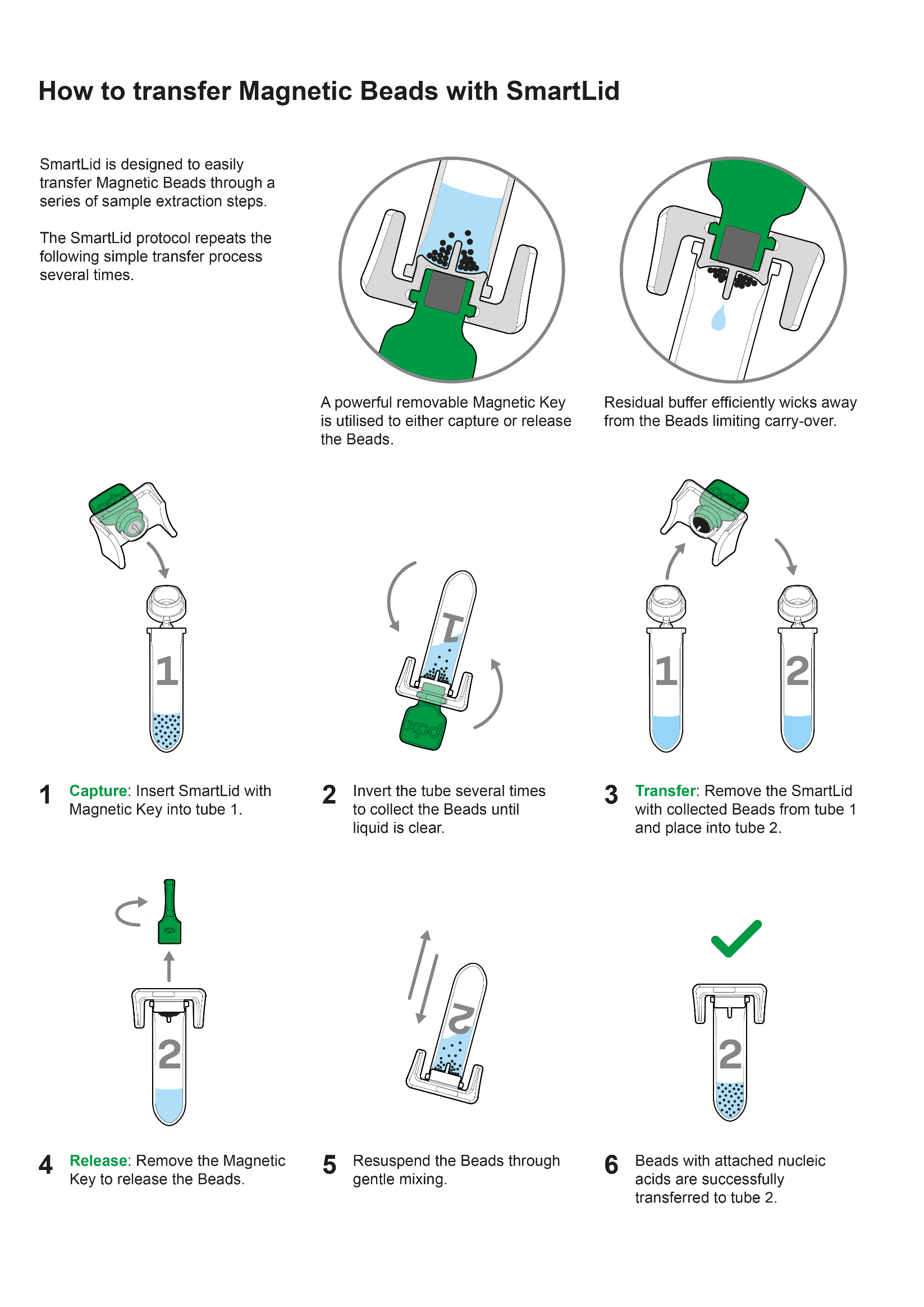

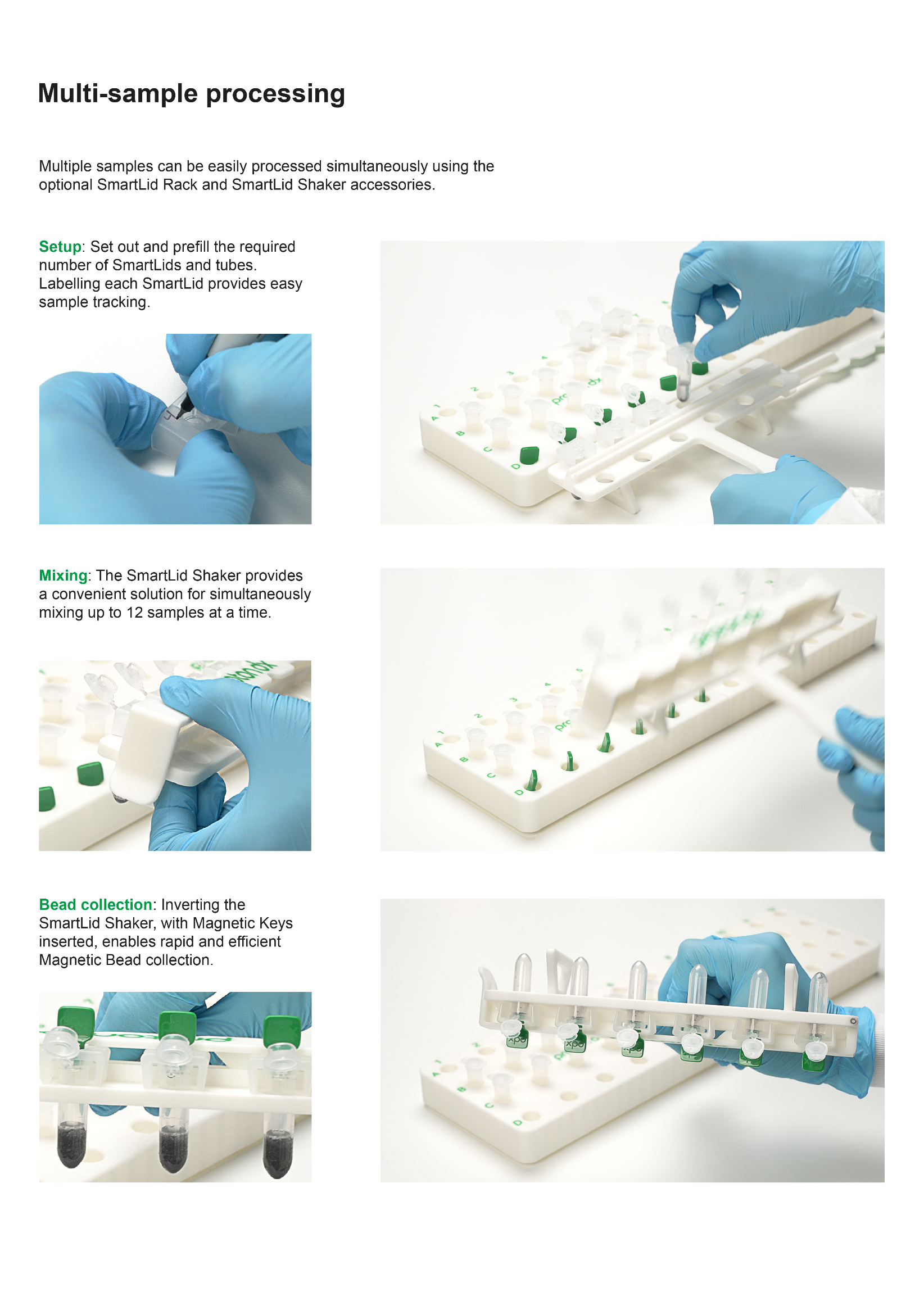

**Supplemental Method 2. Monoplex one-step TaqMan RT-qPCR for influenza subtyping:**

Influenza A(H1N1)pdm09, influenza A(H3), and influenza B/Victoria were analysed using separate target-specific monoplex one-step TaqMan RT-qPCR reactions. Influenza A(H1N1)pdm09 and A(H3) subtyping was performed using the CDC Influenza Virus Real-Time RT-PCR Influenza A(H3/H1pdm09) Subtyping Panel, version 4, for research use only (catalogue number FluRUO-16, 1,000 reactions; International Reagent Resource reference FR-1890; manufactured by the CDC Influenza Division, Atlanta, GA, USA). Influenza B lineage identification was performed using the CDC Influenza B Lineage Genotyping Panel, version 1.1, for research use only (catalogue number FluRUO-11; International Reagent Resource reference FR-1635), supplied through the International Reagent Resource. This panel was used to identify influenza B/Victoria lineage viruses. The WHO reagent listing confirms FluRUO-11/FR-1635 as the CDC Influenza B Lineage Genotyping Panel, version 1.1.

Reactions were prepared using the SuperScript™ III Platinum™ One-Step qRT-PCR Kit with ROX (Invitrogen, catalogue number 11745500). Each reaction had a final volume of 25 µL and contained 12.5 µL of 2× Reaction Mix with ROX, 0.5 µL of SuperScript™ III RT/Platinum™ Taq Mix, 0.5 µL each of the relevant target-specific forward primer, reverse primer, and probe, 5.5 µL of DEPC-treated water, and 5 µL of extracted RNA template. The forward and reverse primers were each used at a final concentration of 800 nM, and the probe was used at a final concentration of 200 nM. Probes were labelled with 6-FAM as the reporter dye and a non-fluorescent quencher.

Amplification was performed using a LightCycler® 480 II instrument (Roche). Reverse transcription was conducted at 50°C for 30 minutes, followed by initial denaturation and enzyme activation at 95°C for 2 minutes. Amplification comprised 45 cycles of denaturation at 95°C for 15 seconds and annealing/extension at 55°C for 30 seconds. Fluorescence was acquired in the FAM channel during each annealing/extension step. Identical reaction conditions, fluorescence-acquisition settings, and assay-specific interpretation criteria were applied to eluates generated by both extraction methods.

Supplemental Table S1. RT-qPCR reaction composition

| **Component** | **Volume per 25 µL reaction** | **Final concentration** |
| --- | --- | --- |
| 2× Reaction Mix with ROX | 12.5 µL | 1× |
| SuperScript III RT/Platinum Taq Mix | 0.5 µL | As supplied |
| Target-specific forward primer | 0.5 µL | 800 nM |
| Target-specific reverse primer | 0.5 µL | 800 nM |
| Target-specific 6-FAM-labelled probe | 0.5 µL | 200 nM |
| DEPC-treated water | 5.5 µL | — |
| Extracted RNA template | 5.0 µL | — |
| **Total reaction volume** | **25.0 µL** | — |

Supplemental Table S2. RT-qPCR thermal cycling programme

| **Stage** | **Temperature** | **Time** | **Cycles** | **Fluorescence acquisition** |
| --- | --- | --- | --- | --- |
| Reverse transcription | 50°C | 30 min | 1 | — |
| Initial denaturation and enzyme activation | 95°C | 2 min | 1 | — |
| Denaturation | 95°C | 15 s | 45 | — |
| Annealing/extension | 55°C | 30 s |  | FAM channel |
